# Clinical Validation of a Multi-protein, Serum-based Assay for Disease Activity Assessments in Multiple Sclerosis

**DOI:** 10.1101/2023.02.08.23285438

**Authors:** Tanuja Chitnis, John Foley, Carolina Ionete, Nabil K. El Ayoubi, Shrishti Saxena, Patricia Gaitan-Walsh, Hrishikesh Lokhande, Anu Paul, Fermisk Saleh, Howard Weiner, Jennifer L. Venzie, Ferhan Qureshi, Michael J. Becich, Fatima Rubio da Costa, Victor M. Gehman, Fujun Zhang, Anisha Keshavan, Kian Jalaleddini, Ati Ghoreyshi, Samia J. Khoury

**Affiliations:** Brigham and Women’s Hospital, Harvard Medical School, Boston, MA, USA; Rocky Mountain Multiple Sclerosis Clinic, Salt Lake City, UT, USA; University of Massachusetts Medical School, Worcester, MA, USA; American University of Beirut, Beirut, Lebanon; The Lockwood Group, Stamford, CT, USA; Octave Bioscience, Inc., Menlo Park, CA, USA

## Abstract

**Background and objectives:** An unmet need exists for validated quantitative tools to measure multiple sclerosis (MS) disease activity and progression. We developed a custom immunoassay-based MS disease activity (MSDA) Test incorporating 18 protein concentrations into an algorithm to calculate four Disease Pathway scores (Immunomodulation, Neuroinflammation, Myelin Biology, and Neuroaxonal Integrity) and an overall Disease Activity score. The objective was to clinically validate the MSDA Test based on associations between scores and clinical/radiographic assessments.

**Methods:** Serum samples (N=614) from patients with MS at multiple sites were split into Train (n=426; algorithm development) and Test (n=188; evaluation) subsets. Subsets were stratified by demographics, sample counts per site, and gadolinium-positive (Gd+) lesion counts; age and sex were used to demographically adjust protein concentrations. MSDA Test results were evaluated for potential association with Gd+ lesion presence/absence, new and enlarging (N/E) T2 lesion presence, and active versus stable disease status (composite endpoint combining radiographic and clinical evidence of disease activity).

**Results:** A multi-protein model was developed (trained and cross-validated) using the Train subset. When applied to the Test subset, the model classified the Gd+ lesion presence/absence, N/E T2 lesion presence, and active versus stable disease status assessments with an area under the receiver operating characteristic (AUROC) of 0.781, 0.750, and 0.768, respectively. In each case, the multi-protein model had significantly (bootstrapped, one-sided *p*<0.05) greater AUROC performance when compared with the top-performing, demographically adjusted (by age and sex) single-protein model based on neurofilament light polypeptide chain. Algorithmic score thresholds corresponded to low, moderate, or high levels of disease activity. Based on the Test subset, the diagnostic odds ratios determined that the odds of having ≥1 Gd+ lesions among samples with a moderate/high Disease Activity score were 4.49 times that of a low Disease Activity score. The odds of having ≥2 Gd+ lesions among samples with a high Disease Activity score were 20.99 times that of a low/moderate Disease Activity score.

**Discussion:** The MSDA Test was clinically validated; the multi-protein model had greater performance compared with the top-performing single-protein model. The MSDA Test may serve as a quantitative and objective tool to enhance care for MS.

## INTRODUCTION

Multiple sclerosis (MS) is an autoimmune, chronic, neuroinflammatory disease of the central nervous system,^1, 2^ with a complex disease course and variable symptoms or manifestations.^1^ The clinical course following the first clinical manifestation of MS, or clinically isolated syndrome (CIS), can vary, but most patients transition to having an MS diagnosis.^3, 4^ Approximately 15% of patients have primary progressive MS, which is usually diagnosed following symptom onset, has no periods of remission, and has a worse prognosis compared with other types of MS.^5-7^ More than 85% of patients have relapsing-remitting MS (RRMS),^7-9^ which is characterized by clinical exacerbations, or relapses, followed by periods of clinical remission, or recovery, as inflammation resolves and remyelination occurs.^1, 5, 7, 9, 10^ Most patients with RRMS enter a progressive phase, which presents as accumulating loss of neurological function over time, a result of demyelination, neuroinflammation, accumulation of neuroaxonal damage, and brain atrophy.^11, 12^ These manifestations contribute to a progressively worsening disability, namely, secondary progressive MS, which can occur with or without further relapses.^5, 8, 13^

The heterogenous variations in the clinical disease course of MS have made diagnosis and prognosis difficult.^9, 14^ Although early diagnosis was based primarily on clinical evidence, the 2017 revision of the McDonald criteria for MS diagnosis has been updated to place greater emphasis on radiographic evidence using magnetic resonance imaging (MRI; eg, dissemination or spread over a minimum of two distinct areas of the central nervous system [CNS], including the brain, spinal cord, and optic nerves and at two different time points), as well as the presence of oligoclonal bands in the cerebral spinal fluid (CSF). Dissemination of lesions can be evaluated by gadolinium enhanced or T2-weighted imaging.^15, 16^ Although the McDonald criteria was recently updated to combine clinical manifestations and radiographic imaging,^15^ these criteria do not always accurately predict disease course, activity, progression, recurrence, or treatment response.^13, 17, 18^ To date, there are no validated clinical tests that leverage multiple serum biomarkers to track disease activity or disease progression in patients with MS. As such, there is an unmet need for clinically validated, objective, quantitative tests that can accurately monitor MS disease activity and progression.^14, 19^

A multi-protein, serum-based biomarker assay was developed to quantitatively measure disease activity using protein concentrations of 18 biomarkers in the serum of patients with all types of MS. The custom multi-protein assay panel was developed and analytically validated using the Olink^®^ Proximity Extension Assay (Olink Proteomics, Uppsala, Sweden) technology.^20^ The comprehensive analytical characterization of this MS disease activity (MSDA) Test was described previously. Briefly, 18 proteins were selected for inclusion into the panel based on results from previously performed research and development studies and incorporated into a final algorithm for calculating four Disease Pathway scores (Immunomodulation, Neuroinflammation, Myelin Biology, and Neuroaxonal Integrity) scores and an overall Disease Activity score (**Supplementary Table 1**).^21^ The objective of the study was to clinically validate the MSDA Test by evaluating the associations of the overall Disease Activity score and the four Disease Pathway scores with gadolinium-positive (Gd+) lesions, new and enlarging T2 (N/E T2) lesions, and active or stable disease status. MS disease status was a combination assessment of Gd+ lesions, N/E T2 lesions, and clinical relapse status.

## METHODS

### Patient samples

A total of 614 serum samples were included from two different sources: 448 retrospective samples from the Serially Unified Multicenter MS Investigation [SUMMIT] consortium; 166 prospective samples from the Rocky Mountain Multiple Sclerosis Clinic (RMMSC). SUMMIT samples were sent to Octave Bioscience, Inc. from three independent sites and studies: Comprehensive Longitudinal Investigation of Multiple Sclerosis at the Brigham and Women’s Hospital (CLIMB; n=195), American University of Beirut Medical Center Study (AMIR; n=202), and University of Massachusetts MS Center—Family Study of Demyelinating Disease (FSDD; n=51). Patient samples selected for this study were intentionally enriched for the presence of Gd+ lesions compared with the general MS population.

SUMMIT was an international multicenter, prospectively enrolled MS cohort study with standardized data structure and analysis groups that can be stratified by demographics, clinical measures, disease relapses, MRI measures, or blood sampling.^22^ Samples from RMMSC were collected as part of the matched serum and MRI for the Disease Activity Test Development Study. Cross-sectional (samples from a single time point) and longitudinal samples (samples from multiple time points from the same patient) from both SUMMIT and RMMSC were included in the analysis. Serum specimens were collected using standard venipuncture and processing protocols. Samples were transferred to Octave Bioscience, Inc. for analysis and stored at −65□C.

### Clinical and radiographic data for biostatistical analysis

An annotated data set containing clinical and radiographical data was obtained for each serum sample. Individual data sets with additional demographic, clinical, and radiographic fields were collected; a combined data set was matched and adjusted so that the annotations were consistently labeled across the study sites and stratified, meaning that the data were split into the Train and Test subsets with a balanced distribution across demographic characteristics, sample counts per site, and Gd+ lesion counts.

### Bioanalytical analysis

All serum samples were analyzed as part of a single experiment performed over several days. Assay plates contained up to 72 samples analyzed in a single well each; four serum controls, three calibrators, and a blank control, were described previously^21^ and assayed in triplicate. Analytical runs were stratified independently of the clinical Train/Test randomization, ensuring that assay plates had balanced distribution across sites, demographics, and presence/absence of or number of Gd+ lesions. Analysis was performed cross-sectionally, using the protein concentrations as predictors and algorithmic features.

Pre-processing and quality control was performed using the Olink^®^ Normalized Protein Expression (NPX) Manager software (Olink Proteomics, Uppsala, Sweden). Built-in quality control (three internal controls that were added into all samples and the external controls) enabled control over the technical performance of the assay at each step of the analysis. These internal controls consisted of an incubation control, extension control, and a detection control. Quality control was performed per assay run and for individual samples at each step of the analysis. Standard deviations for each internal control were established to be below a predetermined threshold (ie, 0.2 NPX) for the entire plate. Median values were calculated for the incubation and detection controls, respectively, for a sample plate. The result of each internal control was required to be within ±0.3 NPX from the plate median. If any of these internal controls deviated from this range, the sample failed quality control and was reanalyzed.

External controls consisted of serum pools with endogenous protein concentrations established at expected levels. Acceptability of a plate run was based on the percent recovery of the serum pools relative to their expected values (ie, ±3 SD). Individual samples or plates that failed the analytical quality control process were rerun.

### Assessments

The primary and exploratory assessments of the study included evaluation of the association of the multi-protein and single-protein models to the radiographic (Gd+ and N/E T2 lesions) and clinical (active vs stable disease status) assessments. Radiographic annotations were derived from brain MRIs for all patients in the study and from spine MRIs when available. Association of single-protein and multi-protein models with the presence or absence of Gd+ lesions, as determined on a matched MRI performed within 60 days of the blood draw, was considered the primary endpoint. Next, the final models were evaluated relative to additional disease activity exploratory assessments (N/E T2 lesions and active vs stable disease status).

Samples were considered active if any Gd+ lesion was present, if any N/E T2 lesion was present, or if there was evidence of a clinical relapse within 30 days. Samples were otherwise considered stable (including samples missing N/E T2 lesion/clinical relapse data). This approach was used to further refine the overall Disease Activity and Disease Pathway algorithms (eg, Immunomodulation, Neuroinflammation, Myelin Biology, and Neuroaxonal Integrity). Association of the results from multi-protein and single-protein modeling with disease progression was an additional exploratory assessment, which is not reported in this manuscript. Classification models were fit to the data with proteomic results as the independent variables and presence or absence of Gd+ lesions as the dependent variable.

### Statistical analysis

The study hypothesis was that a multi-protein model would significantly associate with clinical and radiographic disease activity endpoints and be superior to the highest performing, single-protein model based on the protein biomarkers included in the multi-protein model.

Inclusion criteria for the study required that information for the primary endpoint reference standard (eg, presence or absence of Gd+ lesions) was available for all patient samples. Upon completion of analytical quality control processes and any necessary reanalysis, index test data (eg, protein concentrations) were available for each sample. For secondary and exploratory endpoints, samples with missing data were excluded from the statistical analysis.

The entire data set (N=614) was split into Train and Test subsets. The Train subset was designed to optimize algorithms and included 70% of the total available samples; the Test subset established the performance specifications of the MSDA Test and comprised 30% of the total available samples. The subsets were stratified to ensure a balanced distribution across demographic characteristics that included age, sex, and disease duration; sample counts per site; and Gd+ lesion counts. Analysis was performed in the Train subset to assess unexpected differences in biomarker ranges resulting from preanalytical processing at the four sites from which the samples were obtained.

A two-layer stacked classifier using L2-penalized logistic regression models, which leveraged biological categorizations of biomarkers to calculate Disease Pathway scores, was ultimately developed. A score-based algorithm enabled the four Disease Pathway scores and an overall Disease Activity score to be derived from the probabilities from the Pathway and Disease Activity model and calculated for individual samples (**Figure 1**). Full details on the algorithm and model parameters are presented elsewhere.^23^

**Figure 1.**
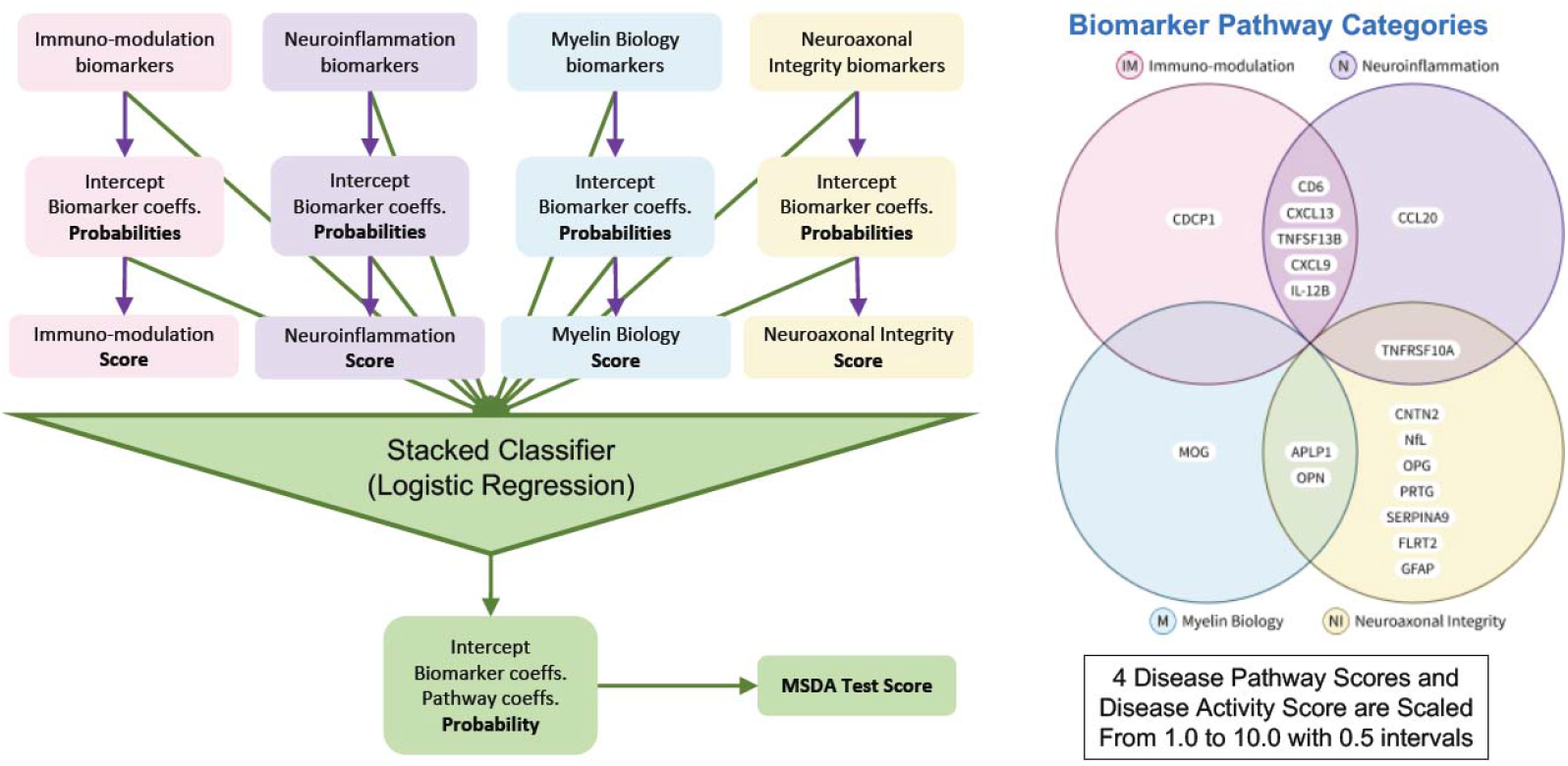
MSDA Test stacked classifier flow chart and biological characterizations model. The first layer of the model consisted of four Disease Pathway algorithms. The second layer of the model utilized the four Disease Pathway probabilities along with the individual age- and sex-adjusted biomarker concentrations as meta-features to determine an overall Disease Activity score that reflected the probability of disease activity. Thresholds were established based on the count of Gd+ lesions for the Disease Activity score, which corresponded to low (1.0–4.0), moderate (4.5–7.0), or high (7.5–10.0) levels of disease activity. MSDA, multiple sclerosis disease activity.

Single-protein models were fit using L2-penalized logistic regression with presence or absence of Gd+ lesions as the dependent variable and an intercept and the protein biomarker as independent variables.

Protein concentrations were limit of quantitation (LOQ)-imputed, log_10_-transformed, and demographically adjusted (with age and sex, based on Ordinary Least Squares [OLS] modeling) prior to being used in the Disease Pathway, Disease Activity, and single-protein models. Sex and age were selected as demographic adjustment variables (if there was a dependence with the protein biomarkers) since they are routinely collected in a clinical setting when blood samples are taken for analysis. Previous research and development studies, samples from a cohort of healthy controls, and those from the Train subset were used to establish the biomarker-specific demographic adjustment strategy, which included removing protein concentration outliers, accounting for OLS coefficient sign consistency across the three studies and establishing statistical significance related to both age and sex.^23^

Metrics for model performance including the area under the receiver operating characteristic (AUROC), sensitivity, specificity, positive predictive value (PPV), negative predictive value (NPV), accuracy, and odds ratios were used to evaluate model performance. The prevalence of Gd+ lesions was enriched in this dataset, and it is important to note that PPV, NPV, and accuracy all depend on the prevalence. An L2-penalization was used to optimize the model and minimize overfitting when training to ensure generalizability to the Test subset. The multi-protein classification model was compared with the top-performing single-protein model (demographically corrected neurofilament light polypeptide chain [NfL] for all disease activity endpoints) to assess the statistical significance of differences in AUROC performance using a boot-strapped (1000 iterations), one-sided test for significance using the pROC^24^ package in R.^25^

The importance of each protein biomarker included in the Disease Activity model was evaluated using mlxtend^26^ by the mean decrease in the AUROC after permutation (repeated 1000 times) of only that protein marker in the trained Pathway and Disease Activity models, compared with the AUROC for predictions with no permutation. Proteins that displayed a larger decrease in the AUROC were more important for the Disease Activity model. NfL was identified as the most important biomarker, followed by tumor necrosis factor superfamily member 13B (TNFSF13B) (**Supplementary Figure 1**).

Statistical analysis was performed to characterize the algorithm at the model level (eg, prior to scoring individual samples) and at the score level. Thresholds were established for the score levels that corresponded to Disease Activity scores based on the number of Gd+ lesions. Patients with no lesions were considered to have low (1.0–4.0) disease activity, patients with 1 Gd+ lesion had moderate (4.5–7.0) disease activity, and patients with ≥2 Gd+ lesions had high (7.5–10.0) disease activity. The low versus moderate/high threshold was selected based on sensitivity. The rationale for selecting sensitivity for the low versus moderate/high threshold was that the presence of any number of Gd+ lesion(s) was an accurate and reliable indicator of active disease. The low/moderate versus high threshold was selected based on accuracy. The rationale for selecting accuracy for the low/moderate versus high threshold was that all samples within this score range were expected to have radiographic evidence of disease activity; the primary factor would therefore be optimizing the ability to distinguish a single lesion versus ≥2 Gd+ lesions.

Statistical analysis for the Train subset was performed by the Octave Bioscience Data Science Team who remained blinded to the analytical results (eg, protein concentrations) and clinical assessments in the Test subset until the algorithm was finalized. To investigate generalizability of the Disease Activity and Pathway algorithms determined using the Train subset, samples from earlier research and development studies were also evaluated prior to the Test subset analysis (analyses not reported here).

### Standard protocol approvals, registration, and patient consents

The study was approved by the Mass General Brigham institutional review board (Somerville, MA, USA) and the WCG institutional review board (Puyallup, WA, USA). All patients provided written informed consent.

### Data availability

Access to anonymized data not published within this article and the study protocol can be made available by request from any qualified investigator once a data-sharing agreement is in place.

## RESULTS

### Demographic and patient characteristics

A total of 614 serum samples were included from two different sources and split into Train (n=426; algorithm development) and Test (n=188; evaluation) subsets. Patient demographics and characteristics were well balanced between Train and Test subsets (**Table 1**).

**Table 1.** Demographic and patient characteristics for the Train and Test subsets and the entire dataset.

| Characteristic* | Train<br>(n=426) | Test<br>(n=188) | Entire<br>(N=614) |
| --- | --- | --- | --- |
| Female | 298 (70.0) | 134 (71.3) | 432 (70.4) |
| Age, years, mean (SD) | 41.6 (12.7) | 42.5 (13.5) | 41.9 (12.9) |
| Disease duration, months, mean (SD) | 9.5 (8.6) | 9.2 (8.8) | 9.4 (8.6) |
| Gd+ lesions |  |  |  |
| 0 | 267 (62.7) | 112 (59.6) | 379 (61.7) |
| 1 | 102 (23.9) | 55 (29.3) | 157 (25.6) |
| ≥2 | 57 (13.4) | 21 (11.2) | 78 (12.7) |
| N/E T2 lesions | 126 (29.6) | 52 (27.7) | 178 (29.0) |
| Disease status† |  |  |  |
| Stable | 251 (58.9) | 104 (55.3) | 355 (57.8) |
| Active | 175 (41.1) | 84 (44.7) | 259 (42.2) |
| Study site |  |  |  |
| American University of Beirut | 143 (33.6) | 59 (31.4) | 202 (32.9) |
| Brigham and Women's Hospital | 134 (31.5) | 61 (32.4) | 195 (31.8) |
| Rocky Mountain MS Clinic | 114 (26.8) | 52 (27.7) | 166 (27.0) |
| University of Massachusetts MS Center | 35 (8.2) | 16 (8.5) | 51 (8.3) |

\*Characteristics were evaluated as n (%) unless otherwise noted. †Active versus stable disease status was defined as a composite endpoint that combined radiographic and clinical evidence of disease activity. Gd+ = gadolinium-positive; MS = multiple sclerosis. N/E = new/enlarging.

### Single-protein model evaluation

The AUROC of demographically corrected, log_10_-transformed, LOQ-imputed individual biomarkers evaluated in the Train and Test subsets ranged from 0.436 to 0.726. NfL was the highest performing protein, with an AUROC of 0.726 for the Test subset. The biomarkers that correlated with nominal significance (*p*<0.05, no multiple hypothesis testing correction) with Gd+ lesion presence included NfL, cluster of differentiation 6 (CD6), C-X-C motif chemokine ligand 13 (CXCL13), interleukin 12β (IL-12β), and TNFSF13B (**Supplementary Table 2)**.

### Multi-protein model performance and optimization

Prior to incorporation of the scoring algorithm, a threshold for the multi-protein stacked classifier was chosen to provide a sensitivity of at least 0.80 for the Train subset. This threshold led to a sensitivity of 0.684 and a specificity of 0.714 in the Test subset. The AUROC of the multi-protein stacked classifier was 0.807 and 0.781 for the Train and Test subsets, respectively, based on Gd+ lesion presence (**Supplementary Table 3)**.

### Clinical validation of the multi-protein model

The multi-protein model developed on the Gd+ lesion presence endpoint using the Train subset was then applied to the Test subset. The final model performance achieved an AUROC of 0.781 relative to the Gd+ lesion presence/absence endpoint, 0.750 relative to the N/E T2 lesion presence endpoint, and 0.768 relative to the active/stable disease endpoint. In each case, the multi-protein model was found to have significantly greater (*p*<0.05) performance when compared with the top-performing single-protein model based on demographically corrected NfL (p<0.05).

By comparison, the single-protein model for demographically corrected NfL had an AUROC of 0.726 for Gd+ lesion presence, 0.660 for N/E T2 lesion presence, and 0.683 for active/stable disease. The multi-protein model also outperformed NfL models based on log_10_-transformed, LOQ-imputed NfL protein concentrations with no demographic correction. The AUROCs for the single-protein, demographically uncorrected NfL models were 0.694, 0.619, and 0.645 for Gd+ lesion presence, N/E T2 lesion presence, and active versus stable disease, respectively (**Figure 2**).

**Figure 2.**
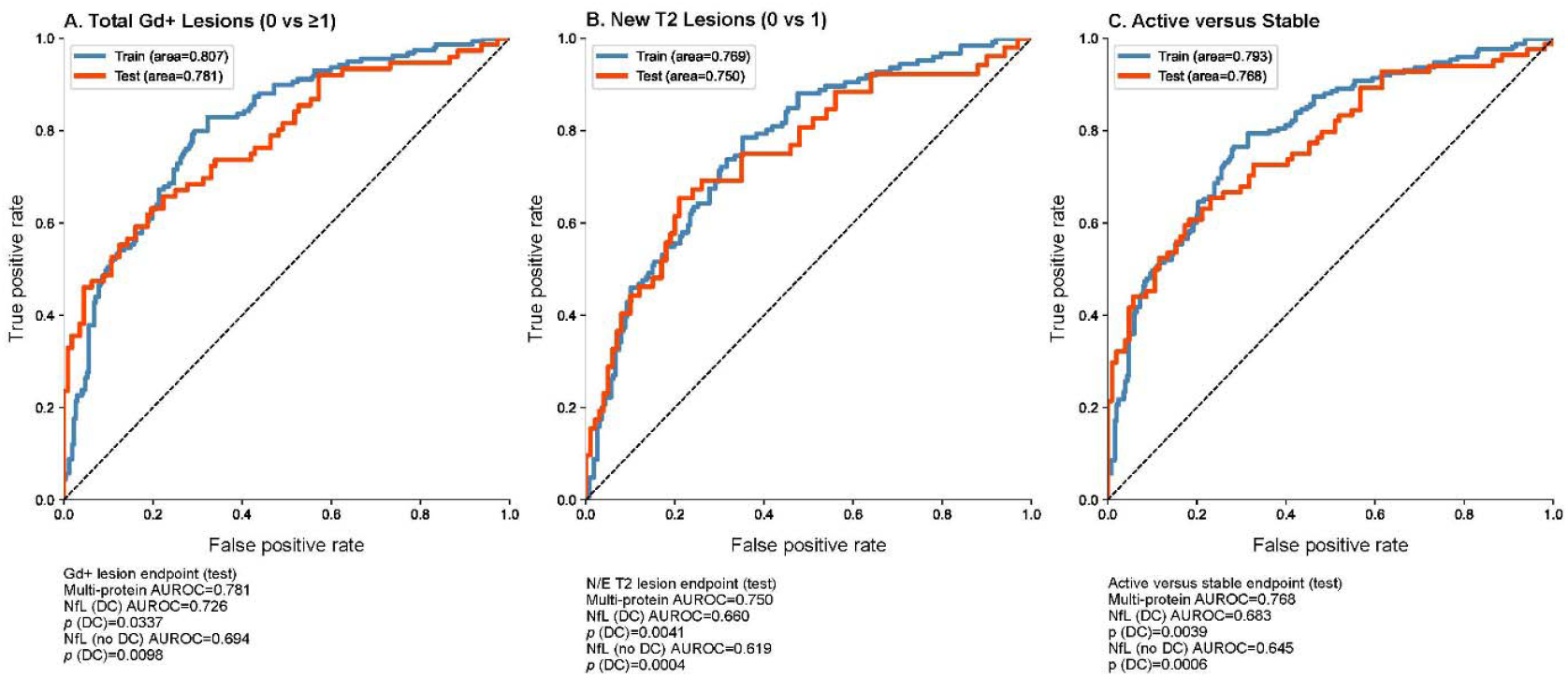
Summary of final performance of the multi-protein model and top performing single-protein demographically corrected (NfL) model in AUROC based on primary (A: Gd+ lesion presence) and exploratory assessments (B: N/E T2 lesion presence and C: active/stable disease status). *P*-values were from a boot-strapped (1000 iterations), one-sided test for significance comparing the multi-protein and single-protein models. AUROC performance and *p*-values are also shown for a single-protein NfL model with no demographic correction. AUROC = area under the receiver operating characteristic curve; DC = demographic correction; Gd+ = gadolinium-positive; N/E = new/enlarging; NfL = neurofilament light polypeptide chain.

Once the final model was optimized, score-level performance of the model in the Train and Test subsets was evaluated using 2 × 2 confusion matrices. Score-level precision for the Train subset is shown in **Table 2**. Selection of the low versus moderate/high threshold was based on sensitivity (for the comparison of no lesions vs ≥1 Gd+ lesions); selection of the low/medium versus high threshold was based on accuracy (for the comparison of 0 or 1 vs ≥2 Gd+ lesions).

**Table 2.** Score level evaluation of the model in the Train and Test subsets by number of Gd+ lesions.

| MS samples | No Gd+ | ≥1 Gd+ | Sensitivity | Specificity | PPV | NPV | Accuracy | Odds Ratio |
| --- | --- | --- | --- | --- | --- | --- | --- | --- |
| Train (n=426) |  |  |  |  |  |  |  |  |
| Low | 150 | 20 | 0.874 | 0.562 | 0.543 | 0.882 | 0.678 | 8.91 |
| Moderate/high | 117 | 139 |  |  |  |  |  |  |
| Test (n=188) |  |  |  |  |  |  |  |  |
| Low | 69 | 20 | 0.737 | 0.616 | 0.566 | 0.775 | 0.665 | 4.49 |
| Moderate/high | 43 | 56 |  |  |  |  |  |  |
| MS samples | 0 or 1 Gd+ | ≥2 Gd+ | Sensitivity | Specificity | PPV | NPV | Accuracy | Odds Ratio |
| Train (n=426) |  |  |  |  |  |  |  |  |
| Low/moderate | 326 | 25 | 0.561 | 0.883 | 0.427 | 0.929 | 0.84 | 9.7 |
| High | 43 | 32 |  |  |  |  |  |  |
| Test (n=188) |  |  |  |  |  |  |  |  |
| Low/moderate | 155 | 8 | 0.619 | 0.928 | 0.52 | 0.951 | 0.894 | 20.99 |
| High | 12 | 13 |  |  |  |  |  |  |

Low, 1.0–4.0; low/moderate, 1.0–7.0; moderate/high, 4.5–10.0; high, 7.5–10. Gd+ = gadolinium-positive; MS = multiple sclerosis; NPV = negative predictive value; PPV = positive predictive value.

For Test samples with ≥1 versus 0 Gd+ lesions, the sensitivity and NPV for the low versus moderate/high cutoff were determined to be 0.737 and 0.775, respectively. A diagnostic odds ratio demonstrated that the odds of having ≥1 Gd+ lesions among samples with a moderate/high Disease Activity score was 4.49 times the odds of having ≥1 Gd+ lesions among samples with a low Disease Activity score (**Table 2**). A comparison of the performance of the Disease Activity score in these samples using the multi-protein model with that of the highest performing single-protein model based on demographically corrected NfL demonstrated that the multi-protein demographically corrected model outperformed the single-protein demographically corrected NfL model (**Figure 3)**.

**Figure 3.**
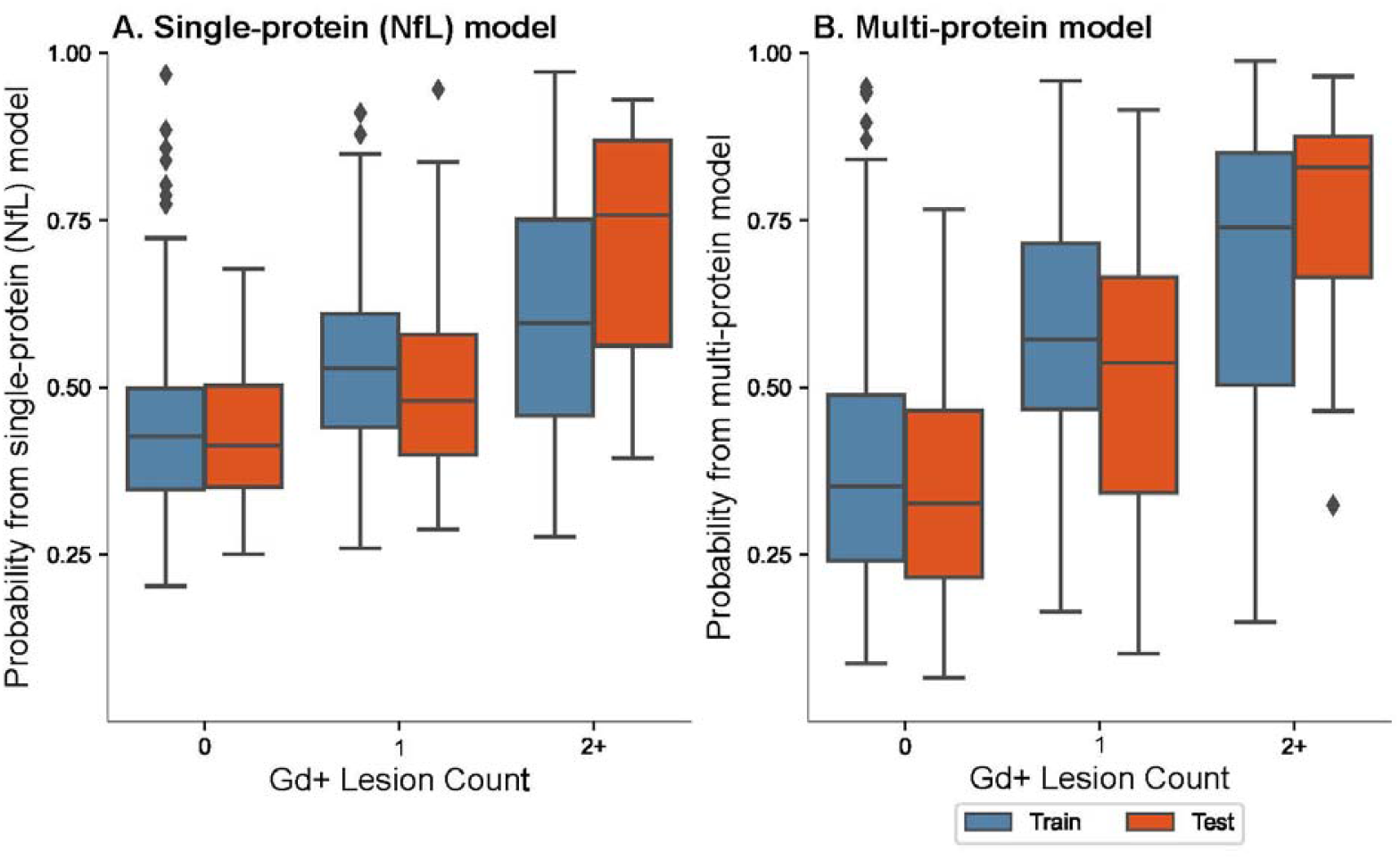
Analysis of disease activity using the A) top-performing single-protein demographically corrected (NfL) model and B) multi-protein model in patients with ≥1 Gd+ lesions. Orange bars = Test subset; Blue bars = Train subset. Gd+ = gadolinium-positive; NfL = neurofilament light polypeptide chain.

For the Test samples with either 0 or 1 Gd+ lesions when compared with those samples with ≥2 Gd+ lesions, the accuracy at the Disease Activity score to predict low/moderate versus high cutoff was determined to be 0.894. The diagnostic odds ratio demonstrated that the odds of having ≥2 Gd+ lesions among samples with a high Disease Activity score were 20.99 times the odds of having ≥2 Gd+ lesions among samples with a low/moderate Disease Activity score (**Table 2**).

The overall performance of the Disease Activity score and four Disease Pathway scores is shown in **Figure 4**. The score distribution and respective box plots with Gd+ lesions for each of the four Disease Pathway scores are shown in **Supplementary Figure 2**. The centering and scaling strategy for the four Disease Pathway scores resulted in sufficient correlation to the overall Disease Activity score while retaining an independent signal. Finally, a stacked bar plot of the results for the Train and Test subsets demonstrated that the calculated Disease Activity score reflected both the likelihood and severity of radiographic disease activity, based on the presence or absence of and count of Gd+ lesions. As shown, patients without Gd+ lesions had a low Disease Activity score (in blue), patients with high Gd+ lesions had a high Disease Activity score (in orange) and those with moderate disease activity had a medium range of Disease Activity score (in yellow; **Figure 5**).

**Figure 4.**
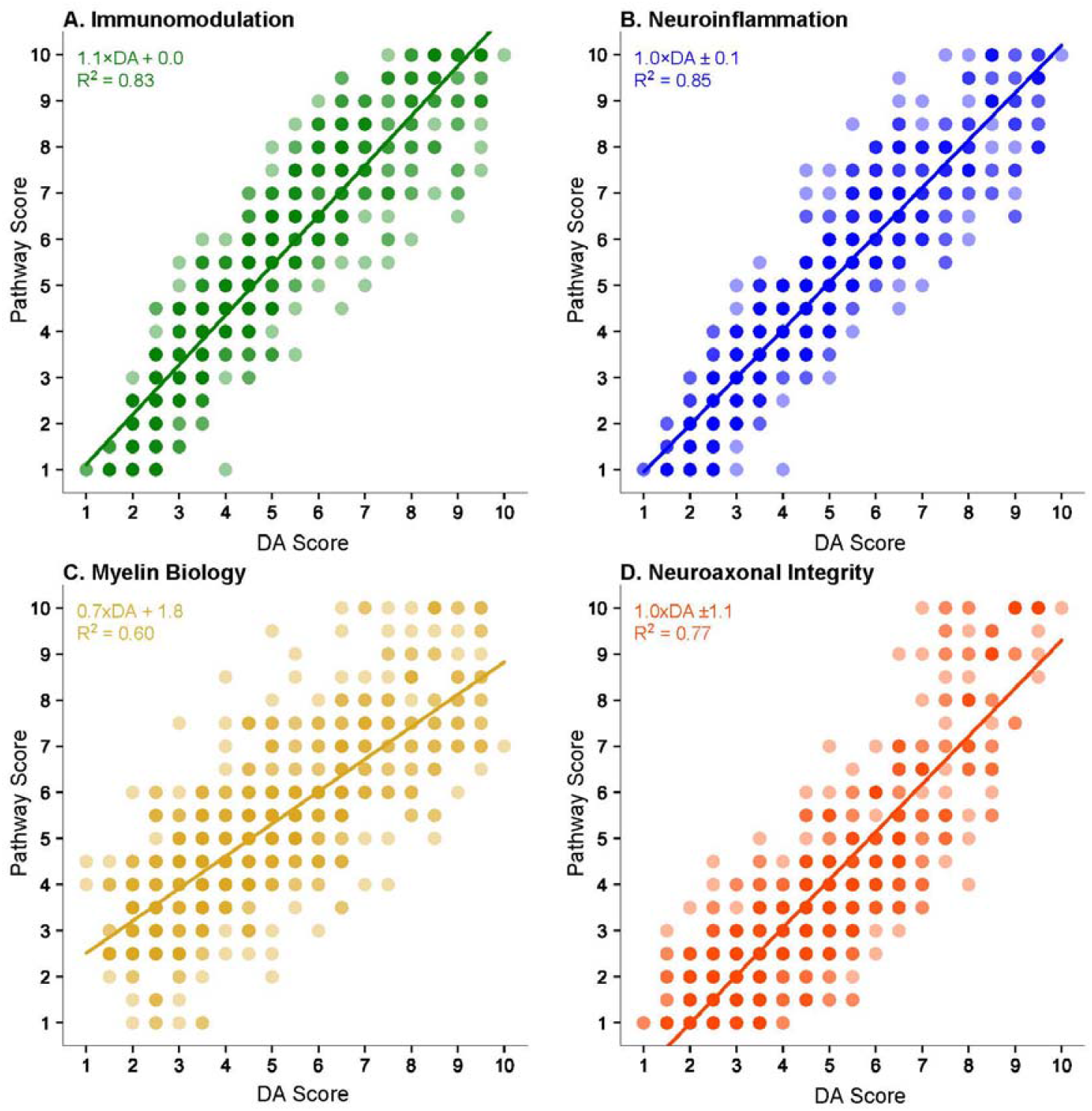
Correlation of the Disease Activity and Disease Pathway (A: Immunomodulation, B: Neuroinflammation, C: Myelin Biology, and D: Neuroaxonal Integrity) scores in the multi-protein model. The solid line indicates the linear regression fit between the Disease Activity and Disease Pathway scores, and the equations for the lines are given in the upper left corner of each figure. DA = Disease Activity; R^2^ = coefficient of determination.

**Figure 5.**
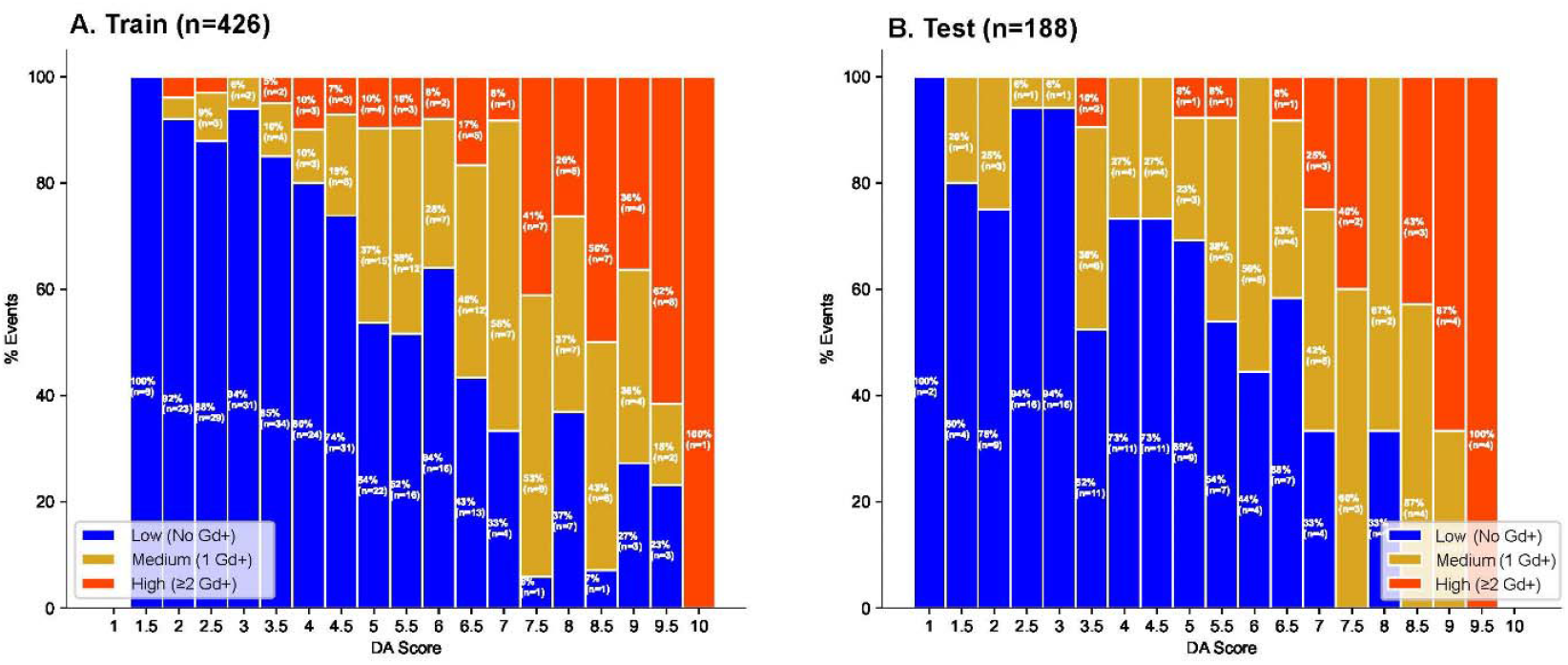
Stacked bar plots of the Disease Activity score in the A) Train and B) Test subsets. Low disease activity (blue) was associated with no Gd+ lesions, moderate disease activity (yellow) was associated with ≥1 Gd+ lesions, and high disease activity (orange) was associated with ≥2 Gd+ lesions. Bars for Disease Activity scores for which there were no samples are blank. DA = Disease Activity; Gd+ = gadolinium-positive.

The distribution of individual biomarkers, and the Disease Activity and Disease Pathway scores obtained from the MSDA Test were analyzed for each of six disease-modifying therapy (DMT) categories (anti-CD20s, natalizumab, interferons, dimethyl fumarate, fingolimod, and glatiramer acetate) in 466 samples, reflecting the therapy the patient was on at the time of the blood draw. The remaining samples did not fall in one of these six categories and were categorized as either other (treated with a DMT that was not analyzed independently due to low sample count) or blank (information relating to therapy was not provided in the clinical dataset). Significant analysis of variance (ANOVA) Bonferroni corrected *p-*values were observed for 14 of the 18 individual biomarkers utilized in the MSDA Test algorithm (**Supplementary Figure 3**). Samples (n=40) from the anti-CD20 category had the lowest Disease Activity score on average (3.11 ± 1.77), followed by samples (n=129) from the natalizumab category (4.17 ± 1.63). The highest Disease Activity score on average was associated with samples (n=62) from the glatiramer acetate category (6.39 ± 1.67; **Supplementary Table 4**).

### Classification of evidence

The MSDA Test is a multi-protein, serum-based biomarker assay designed to quantitatively measure disease activity using the protein levels of biomarkers present in the serum of patients with MS. Protein concentrations were LOQ imputed, log_10_ transformed, and demographically adjusted for age and sex. The combination of multiple proteins was used to calculate four Disease Pathway scores and an overall Disease Activity score. The protein selection was intended to reflect the various biological pathways associated with MS pathophysiology. Using associations of the results from the multi-protein and single-protein models and the radiographic/clinical assessments, the MSDA Test was clinically validated in this study. This study provided Class II evidence to demonstrate the clinical validation of MSDA Test for disease activity assessments in MS.

## DISCUSSION

There are currently no validated clinical tests that leverage multiple serum biomarkers to monitor disease activity or progression in patients with MS. We have previously established that the MSDA Test is accurate, sensitive, precise, and robust, which serves as a critical first step in the validation of this assay.^23^ The MSDA Test uses 18 protein biomarkers, which reflect various biological pathways associated with MS pathophysiology.

In this study, we successfully demonstrated the clinical validation of the MSDA Test. All disease activity assessments, namely, Gd+ lesions, N/E T2 lesions, and active/stable disease status, showed association with the Disease Pathway and overall Disease Activity scores from the MSDA Test. The multi-protein model had significantly (bootstrapped, one-sided *p*<0.05) greater performance compared with the top-performing single-protein model based on demographically corrected NfL in all assessments. The MSDA Test performed well when differentiating samples from patients with ≥1 Gd+ lesions versus no lesions, as well as when differentiating samples from patients with ≥2 Gd+ lesions versus 0 or 1 lesions.

The highest-performing single-protein biomarker for all disease activity endpoints was NfL. NfL has been demonstrated to be a prognostic indicator of disease activity in MS.^27-31^ Elevated serum and CSF levels of NfL correlate with neuronal cell damage and brain atrophy.^28, 29, 32-34^ It has been used to predict long-term clinical outcomes of MS^34, 35^ and guide treatment decisions.^28, 29, 34, 36, 37^ However, NfL is not specific for MS and is elevated in several neurodegenerative diseases.^38-40^ Furthermore, the protein is released into CSF and blood as a result of neuroaxonal damage, reflecting pathophysiology downstream of immune-mediated inflammatory pathways.^29, 38, 39^ In single-protein analyses, high-performing biomarkers in addition to NfL in this study included IL-12β, CXCL13, and TNFSF13B, all of which have been found to be potential biomarkers for MS in other studies.^29, 34, 41-46^ Our analysis revealed that the multi-protein model outperformed single-protein models for each of these biomarkers, which indicated a more accurate representation of the various pathways, processes, and cell types involved in a complex disease state, such as MS, by a combination of biomarkers.^20^

The MSDA Test was developed to favor sensitivity over specificity. In biomarker validation studies, there is typically a tradeoff between sensitivity and specificity for assay development.^14^ Development of a highly sensitive model, which can produce a higher degree of false-positive results and have reduced specificity is critical to the identification of patients with subradiographic and subclinical disease activity. Detection of early-stage MS remains challenging when using conventional clinical or radiographic assessments.^17-19^ We believe that the MSDA Test utilizes a well-balanced sensitivity and specificity combination, which can play a key role in the identification of patients with subradiographic and subclinical MS prior to detection of clear clinical or radiographic manifestations. This more sensitive detection will allow for optimal and timely treatment, which can positively impacts patient outcomes.^47^ Another advantage of a blood-based approach is that disease activity can be detected regardless of where in the CNS lesions have occurred. Brain MRIs are more frequently used in the assessment of patients with MS, although spinal lesions are a common occurrence in MS as well.^15, 48^

A disease activity measurement tool should reflect therapeutic efficacy and be characterized relative to the biological impact of various mechanisms of action. In the DMT analysis, the lowest average Disease Activity scores were observed in the anti-CD20 and natalizumab groups, which represent the highest efficacy therapies in our categorizations. Lower Disease Activity scores were observed in patients without radiographic evidence of disease activity (0 Gd+ lesions) and there was a direct correlation between Disease Activity and Disease Pathway scores with Gd+ lesion counts across all DMT categories. Future studies will expand upon the DMT analysis to factor in a patient’s overall disease duration, their duration on the DMT, and their previous DMT history.^49^

Our study has limitations. The patient samples in this study were obtained from four different sites. Practice at different sites, including sample preparation techniques, may introduce potential differences between the data sets. Despite this challenge, the MSDA Test showed successful performance in the test subset, demonstrating its promise of real-world performance.

With the successful clinical validation of the MSDA Test, we envision several potential uses in the future, including a routine surveillance test to better monitor disease activity and progression (eg, distinguish inflammation from silent disease progression), especially in patients considered to have stable disease, and to track new/worsening symptoms, as well as an evaluation test of treatment response, or in consideration of alternative treatment options. We also wish to expand the analysis to investigate the association between overall Disease Activity and Disease Pathway scores and additional assessments (eg, Expanded Disability Status Scale; Patient Determined Disease Steps). Evaluation of the MSDA Test in a larger population of patients with MS in a real-world setting is also valuable. The MSDA Test is intended to complement standard radiographic imaging and clinical assessment and promote individualized disease management.^14, 19, 50^

## GLOSSARY

AMIR: American University of Beirut Medical Center Study
ANOVA: analysis of variance
AUROC: area under the receiver operating characteristic
CD6: cluster of differentiation 6
CIS: clinically isolated syndrome
CLIMB: Comprehensive Longitudinal Investigation of Multiple Sclerosis at the Brigham and Women’s Hospital
CNS: central nervous system
CSF: cerebral spinal fluid
CXCL13: C-X-C motif chemokine ligand 13
DMT: disease-modifying therapy
FSDD: Family Study of Demyelinating Disease
Gd+: gadolinium-positive
IL-12β: interleukin 12β
LOQ: limit of quantitation
MRI: magnetic resonance imaging
MS: multiple sclerosis
MSDA: multiple sclerosis disease activity
N/E: new and enlarging
NfL: neurofilament light polypeptide chain
NPV: negative predictive value
NPX: normalized protein expression
OLS: ordinary least squares
PPV: positive predictive value
RMMSC: Rocky Mountain Multiple Sclerosis Clinic
RRMS: relapsing-remitting multiple sclerosis
SUMMIT: Serially Unified Multicenter Multiple Sclerosis Investigation
TNFSF13B: tumor necrosis factor superfamily member 13B

## ACKNOWLEDGMENTS

Copyediting assistance was provided by The Lockwood Group (Stamford, CT) and was funded by Octave Bioscience, Inc.

### Appendix 1. Author contributions

| Name | Institution and location | Contribution |
| --- | --- | --- |
| Tanuja Chitnis | Brigham and Women's Hospital, Harvard Medical School, Boston, MA | Major role in the acquisition of data; substantial contributions to the conception or design of the work; analysis, or interpretation of data for the work; revising work critically for important intellectual content; final approval of the version to be published; agreement to be accountable for all aspects of the work |
| John Foley | Rocky Mountain Multiple Sclerosis Clinic, Salt Lake City, Utah | Major role in the acquisition of data; analysis, or interpretation of data for the work; revising work critically for important intellectual content; final approval of the version to be published; agreement to be accountable for all aspects of the work |
| Carolina Ionete | University of Massachusetts Medical School, Worcester, MA, USA | Major role in the acquisition of data; analysis, or interpretation of data for the work; revising work critically for important intellectual content; final approval of the version to be published; agreement to be accountable for all aspects of the work |
| Nabil K. El Ayoubi | American University of Beirut, Beirut, Lebanon | Analysis, or interpretation of data for the work; revising work critically for important intellectual content; final approval of the version to be published; agreement to be accountable for all aspects of the work |
| Shrishti Saxena | Brigham and Women's Hospital, Harvard Medical School, Boston, MA | Analysis, or interpretation of data for the work; revising work critically for important intellectual content; final approval of the version to be published; agreement to be accountable for all aspects of the work |
| Patricia Gaitan-Walsh | Brigham and Women's Hospital, Harvard Medical School, Boston, MA | Analysis, or interpretation of data for the work; revising work critically for important intellectual content; final approval of the version to be published; agreement to be accountable for all aspects of the work |
| Hrishikesh Lokhande | Brigham and Women's Hospital, Harvard Medical School, Boston, MA | Analysis, or interpretation of data for the work; revising work critically for important intellectual content; final approval of the version to be published; agreement to be accountable for all aspects of the work |
| Anu Paul | Brigham and Women's Hospital, Harvard Medical School, Boston, MA | Analysis, or interpretation of data for the work; revising work critically for important intellectual content; final approval of the version to be published; agreement to be accountable for all aspects of the work |
| Fermisk Saleh | Brigham and Women's Hospital, Harvard Medical School, Boston, MA | Analysis, or interpretation of data for the work; revising work critically for important intellectual content; final approval of the version to be published; agreement to be accountable for all aspects of the work |
| Howard Weiner | Brigham and Women's Hospital, Harvard Medical School, Boston, MA | Analysis, or interpretation of data for the work; revising work critically for important intellectual content; final approval of the version to be published; agreement to be accountable for all aspects of the work |

| **Name** | **Institution and location** | **Contribution** |
| --- | --- | --- |
| Jennifer L. Venzie | The Lockwood Group,<br>Stamford, CT | Drafting manuscript; final approval of the version to be published; agreement to be accountable for all aspects of the work |
| Ferhan Qureshi | Octave Bioscience, Inc., Menlo<br>Park, CA | Substantial contributions to the conception or design of the work; analysis, or interpretation of data for the work; revising work critically for important intellectual content; final approval of the version to be published; agreement to be accountable for all aspects of the work |
| Michael J. Becich | Octave Bioscience, Inc., Menlo<br>Park, CA | Analysis, or interpretation of data for the work; revising work critically for important intellectual content; final approval of the version to be published; agreement to be accountable for all aspects of the work |
| Fatima Rubio da Costa | Octave Bioscience, Inc., Menlo<br>Park, CA | Analysis, or interpretation of data for the work; revising work critically for important intellectual content; final approval of the version to be published; agreement to be accountable for all aspects of the work |
| Victor M. Gehman | Octave Bioscience, Inc., Menlo<br>Park, CA | Analysis, or interpretation of data for the work; revising work critically for important intellectual content; final approval of the version to be published; agreement to be accountable for all aspects of the work |
| Fujun Zhang | Octave Bioscience, Inc., Menlo<br>Park, CA | Analysis, or interpretation of data for the work; revising work critically for important intellectual content; final approval of the version to be published; agreement to be accountable for all aspects of the work |
| Anisha Keshavan | Octave Bioscience, Inc., Menlo<br>Park, CA | Analysis, or interpretation of data for the work; revising work critically for important intellectual content; final approval of the version to be published; agreement to be accountable for all aspects of the work |
| Kian Jalaleddini | Octave Bioscience, Inc., Menlo<br>Park, CA | Analysis, or interpretation of data for the work; revising work critically for important intellectual content; final approval of the version to be published; agreement to be accountable for all aspects of the work |
| Ati Ghoreyshi | Octave Bioscience, Inc., Menlo<br>Park, CA | Analysis, or interpretation of data for the work; revising work critically for important intellectual content; final approval of the version to be published; agreement to be accountable for all aspects of the work |
| Samia J. Khoury | American University of Beirut,<br>Beirut, Lebanon | Major role in the acquisition of data; analysis, or interpretation of data for the work; revising work critically for important intellectual content; final approval of the version to be published; agreement to be accountable for all aspects of the work |

## Supplementary Materials

**Supplementary Table 1.** List of the 18 proteins selected for assay inclusion.

| **Biomarker** | **Full name (alias)** | **UniProt identifier** | **Biological pathways** |
| --- | --- | --- | --- |
| *APLP1* | Amyloid beta precursor-like protein 1 | P51693 | Myelin Biology, Neuroaxonal Integrity |
| *CCL20* | C-C motif chemokine ligand 20 (MIP 3-alpha) | P78556 | Neuroinflammation |
| *CD6* | Cluster of differentiation 6 | P30203 | Immunomodulation, Neuroinflammation |
| *CDCP1* | CUB domain-containing protein 1 | Q9H5V8 | Immunomodulation |
| *CNTN2* | Contactin 2 | Q02246 | Neuroaxonal Integrity |
| *CXCL13* | Chemokine (C-X-C motif) ligand 13 | P02462 | Immunomodulation, Neuroinflammation |
| *CXCL9* | Chemokine (C-X-C motif) ligand 9 (MIG) | O43927 | Immunomodulation, Neuroinflammation |
| *FLRT2* | Fibronectin leucine-rich repeat transmembrane protein | O43155 | Neuroaxonal Integrity |
| *GFAP* | Glial fibrillary acidic protein | P14136 | Neuroaxonal Integrity |
| *IL-12β* | Interleukin-12 subunit beta | P29460 | Immunomodulation, Neuroinflammation |
| *MOG* | Myelin oligodendrocyte glycoprotein | Q16653 | Myelin Biology |
| *NfL* | Neurofilament light polypeptide chain | P07196 | Neuroaxonal Integrity |
| *OPG* | Osteoprotegerin | O00300 | Neuroaxonal Integrity |
| *OPN* | Osteopontin | P10451 | Myelin Biology, Neuroaxonal Integrity |
| *PRTG* | Protogenin | Q2VWP7 | Neuroaxonal Integrity |
| *SERPINA9* | Serpin family A member 9 | Q86WD7 | Neuroaxonal Integrity |
| *TNFRSF10A* | Tumor necrosis factor receptor superfamily member 10A (TRAIL-R1) | O00220 | Neuroinflammation, Neuroaxonal Integrity |
| *TNFSF13B* | Tumor necrosis factor superfamily member 13B (BAFF) | Q9Y275 | Immunomodulation, Neuroinflammation |

**Supplementary Table 2.** Single-protein model performance of individual biomarker proteins (LOQ-imputed, log_10_ transformed, and demographically corrected for age and sex) based on Gd+ lesion assessments. Nominal *p*-values from a two-sided T-test assuming equal variances between groups (no Gd+ lesions vs ≥1 Gd+ lesions) are reported.

| Protein assay | Train (n=426) |  | Test (n=188) |  |
| --- | --- | --- | --- | --- |
|  | AUROC | *p*-value | AUROC | *p*-value |
| NfL | 0.726 | <0.0001 | 0.726 | <0.0001 |
| MOG | 0.576 | 0.022 | 0.547 | 0.208 |
| CD6 | 0.499 | 0.954 | 0.583 | 0.02 |
| CXCL13 | 0.586 | 0.064 | 0.645 | 0.003 |
| CXCL9 | 0.53 | 0.634 | 0.436 | 0.339 |
| CDCP1 | 0.598 | 0.001 | 0.59 | 0.051 |
| CCL20 | 0.526 | 0.619 | 0.492 | 0.542 |
| OPG | 0.499 | 0.79 | 0.464 | 0.369 |
| IL-12β | 0.619 | <0.0001 | 0.606 | 0.008 |
| APLP1 | 0.522 | 0.623 | 0.536 | 0.502 |
| TNFRSF10A | 0.525 | 0.352 | 0.453 | 0.275 |
| SERPINA9 | 0.505 | 0.92 | 0.529 | 0.412 |
| PRTG | 0.478 | 0.507 | 0.504 | 0.379 |
| FLRT2 | 0.53 | 0.431 | 0.545 | 0.191 |
| TNFSF13B | 0.651 | <0.0001 | 0.603 | 0.003 |
| OPN | 0.525 | 0.578 | 0.479 | 0.306 |
| CNTN2 | 0.531 | 0.414 | 0.534 | 0.538 |
| GFAP | 0.595 | 0.014 | 0.552 | 0.267 |

Green shading indicates a nominal *p*<0.05.

**Supplementary Table 3.** Model-level performance of the multi-protein model in the Train and Test subsets.

|  | **AUROC** | **Accuracy** | **Precision** | **Sensitivity** | **Specificity** | **NPV** | **PPV** | **Odds ratio** |
| --- | --- | --- | --- | --- | --- | --- | --- | --- |
| Train (n=426) | 0.807 | 0.725 | 0.598 | 0.805 | 0.678 | 0.854 | 0.598 | 8.69 |
| Test (n=188) | 0.781 | 0.702 | 0.619 | 0.684 | 0.714 | 0.769 | 0.619 | 5.417 |

Model-level performance was evaluated on the Disease Activity probabilities before the algorithm was used to calculate the overall Disease Activity score.
AUROC = area under the receiver operating characteristic; NPV = negative predictive value; PPV = positive predictive value.

**Supplementary Table 4:**
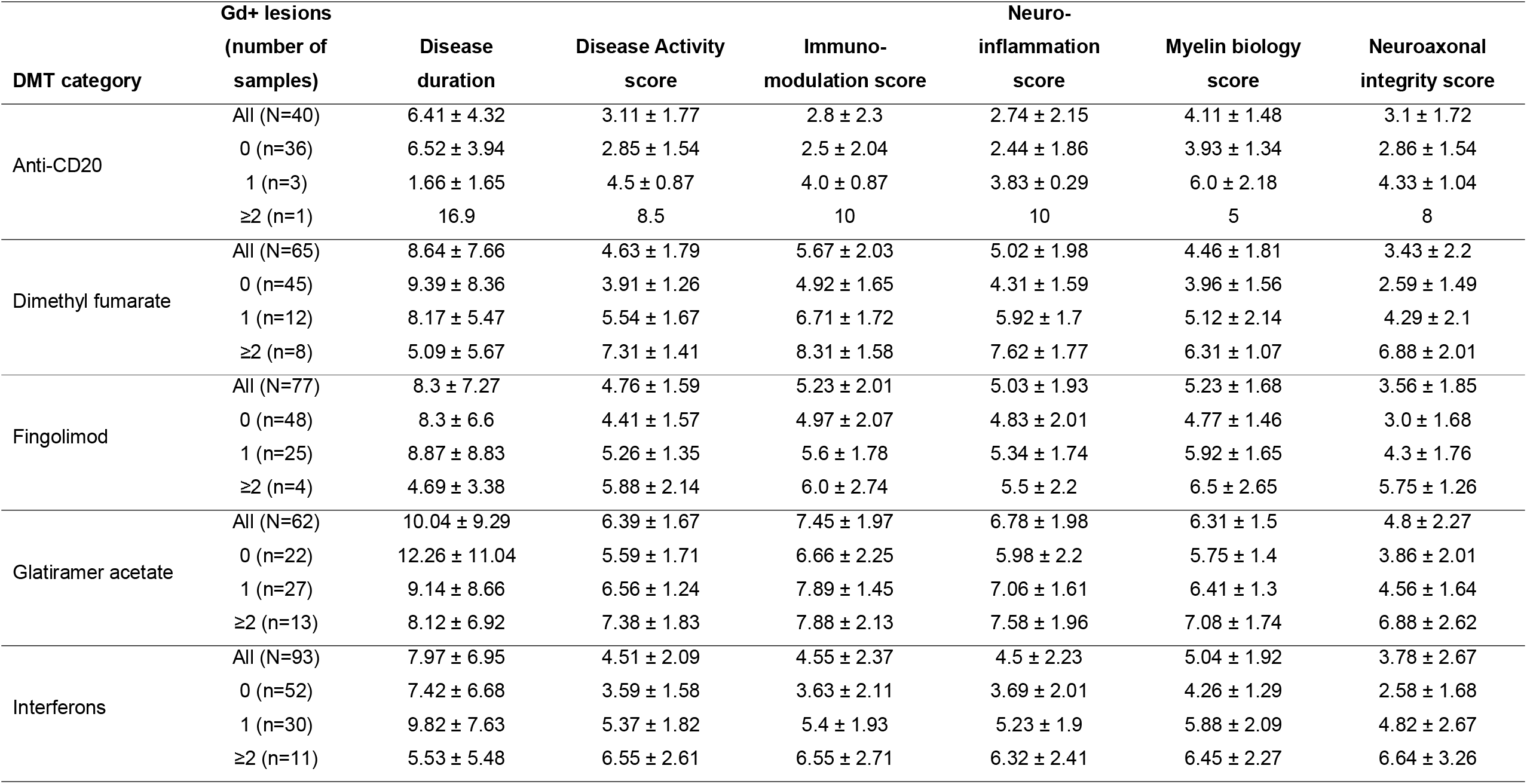
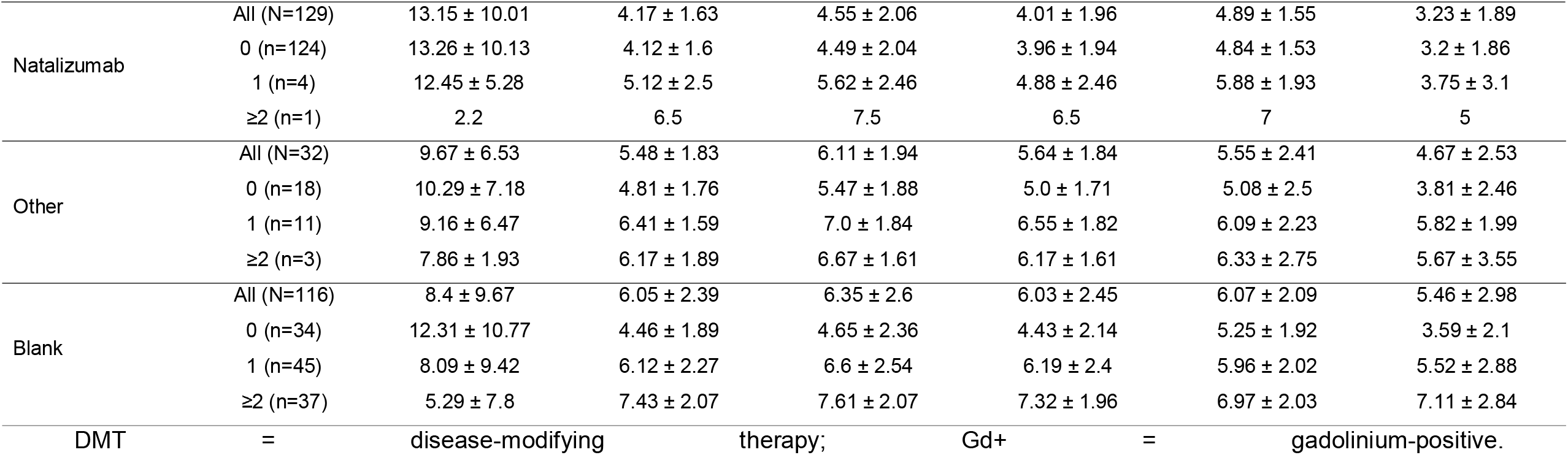
Mean±SDDisease Activity and Disease Pathway scores categorized by disease-modifying therapy.

**Supplementary Figure 1.**
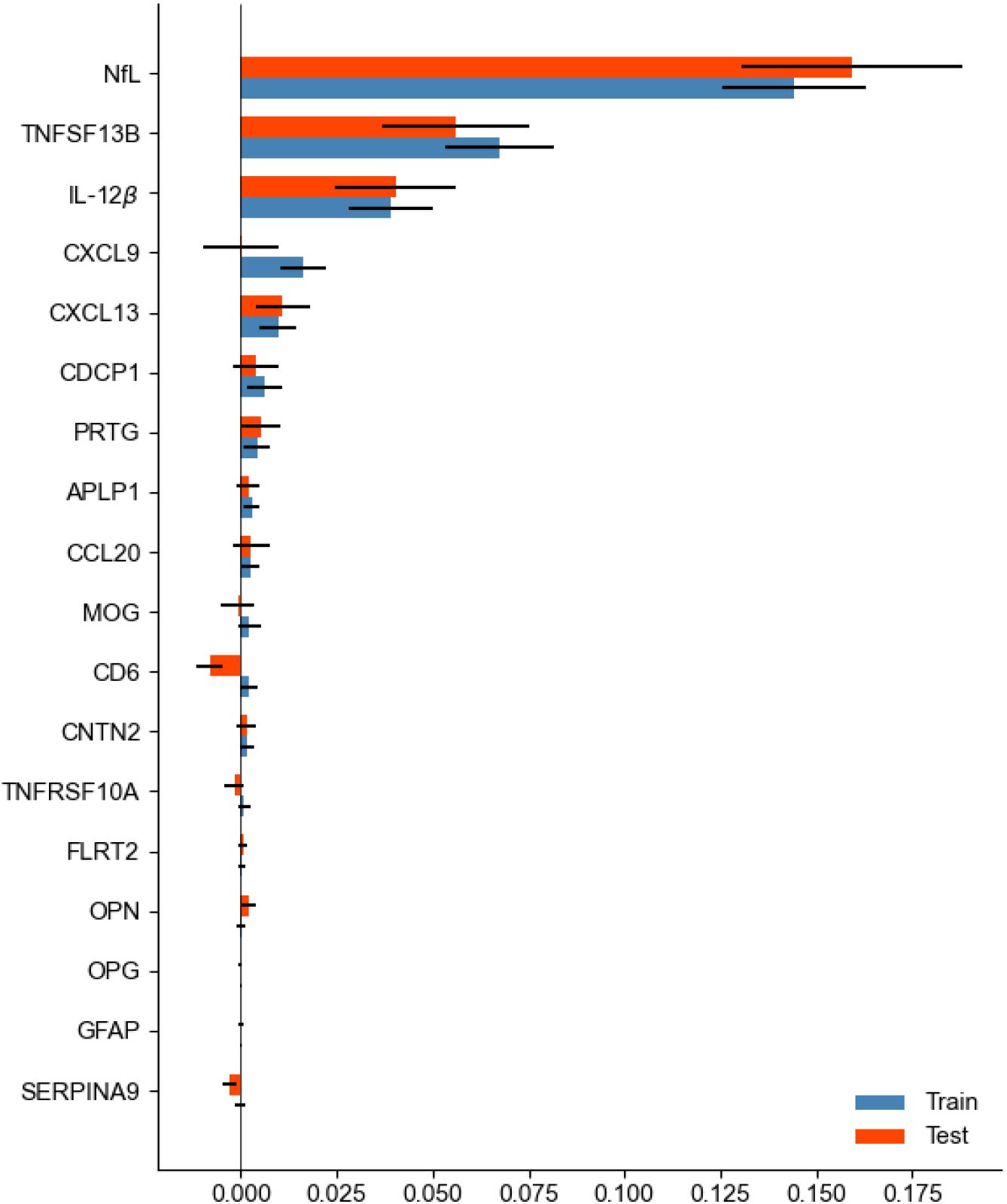
Feature importance demonstrated by mean AUROC decrease (no Gd+ lesions vs ≥1 Gd+ lesions) after permutation (1000 times) for each biomarker in the Disease Activity stacked classifier model and the input Pathway models. Biomarkers with a larger positive decrease in AUROC have greater importance for the Disease Activity model. NfL was identified as the most important feature, followed by TNFSF13B. Error bars correspond to 2.5th and 97.5th percentiles. AUROC = area under the receiver operating characteristic; Gd+ = gadolinium-positive; NfL = neurofilament light polypeptide chain; TNFSF13B = tumor necrosis factor superfamily member 13B.

**Supplementary Figure 2.**
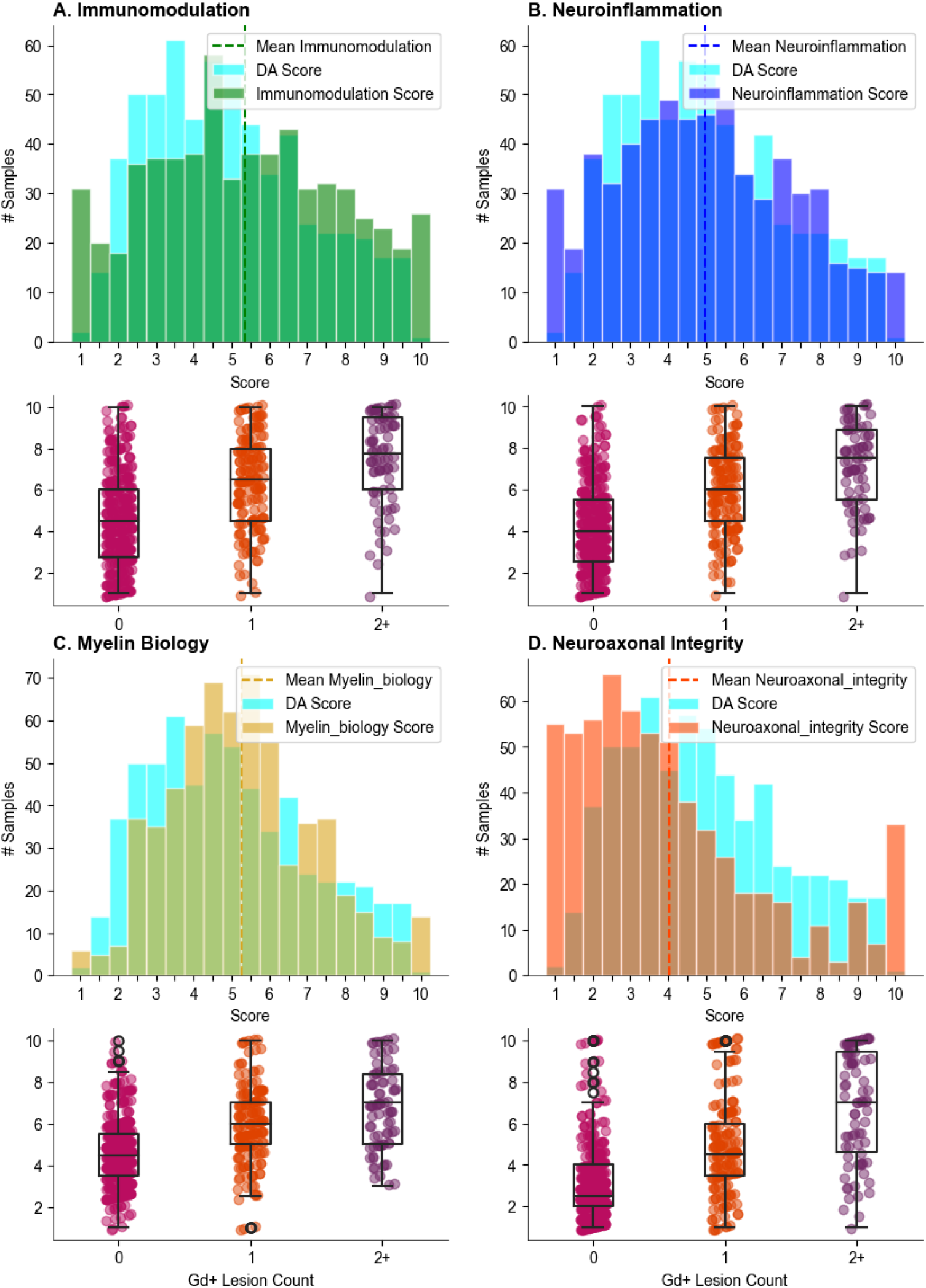
Score distribution and respective box plots with Gd+ lesions for A) Immunomodulation, B) Neuroinflammation, C) Myelin Biology, and D) Neuroaxonal Integrity scores in the multi-protein model. DA = Disease Activity; Gd+ = gadolinium-positive.

**Supplementary Figure 3.**
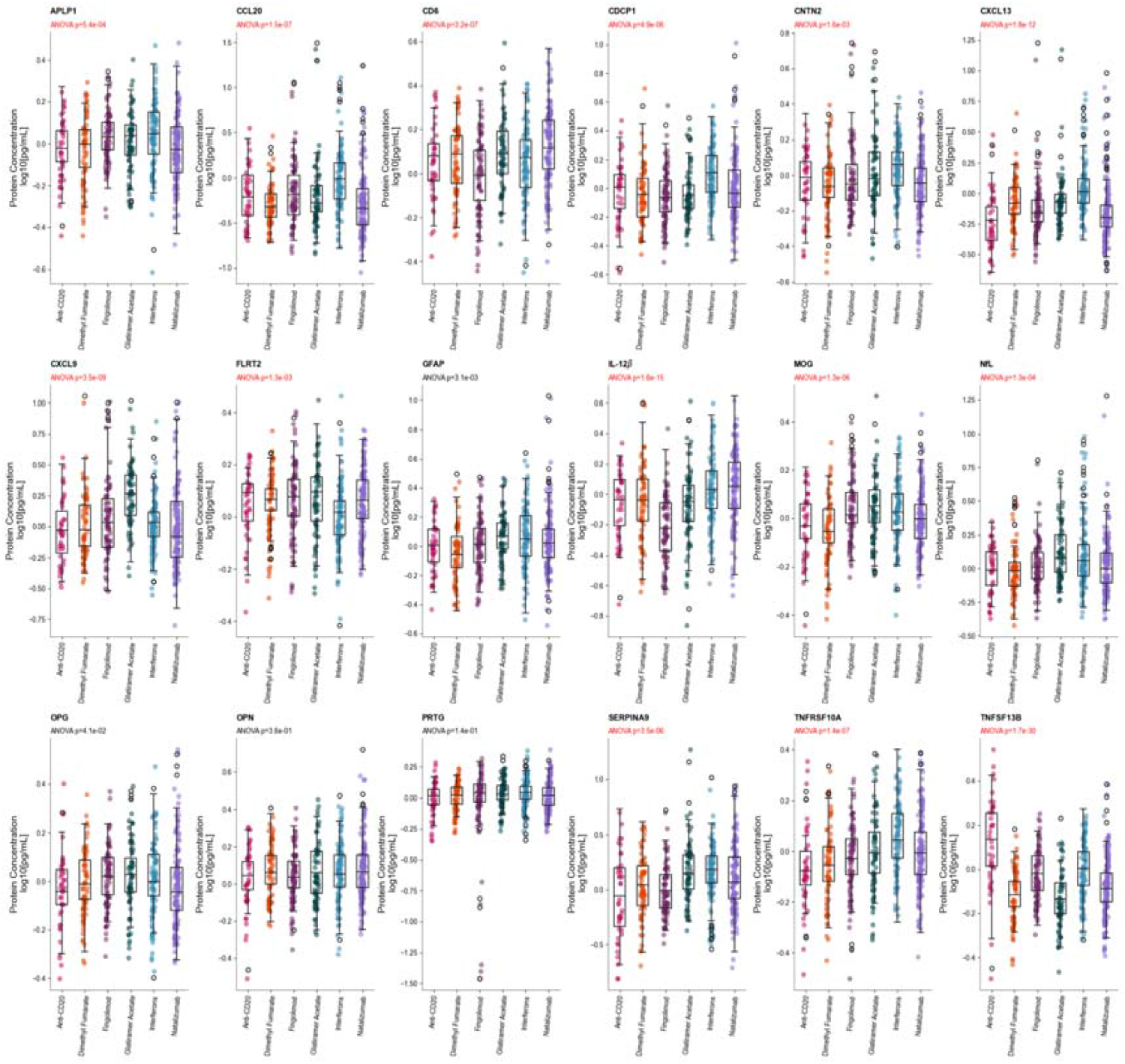
Protein biomarker distributions by disease-modifying therapy class. Significant ANOVA Bonferroni corrected *p*-values, based on biomarkers across DMT categories, are marked in red. Nominal ANOVA *p*-values are reported. ANOVA = analysis of variance; DMT = disease-modifying therapy.

## REFERENCES

1. Compston A, Coles A. Multiple sclerosis. Lancet 2008;372:1502–1517.

2. Weiner HL. Multiple sclerosis is an inflammatory T-cell–mediated autoimmune disease. Arch Neurol 2004;61:1613–1615.

3. Miller DH, Chard DT, Ciccarelli O. Clinically isolated syndromes. Lancet Neurol 2012;11:157–169.

4. Lublin FD, Reingold SC, Cohen JA, et al. Defining the clinical course of multiple sclerosis: the 2013 revisions. Neurology 2014;83:278–286.

5. Lublin FD, Reingold SC. Defining the clinical course of multiple sclerosis: results of an international survey. National Multiple Sclerosis Society (USA) Advisory Committee on Clinical Trials of New Agents in Multiple Sclerosis. Neurology 1996;46:907–911.

6. Cottrell DA, Kremenchutzky M, Rice GPA, et al. The natural history of multiple sclerosis:a geographically based study: 5. The clinical features and natural history of primary progressive multiple sclerosis. Brain 1999;122:625–639.

7. Confavreux C, Vukusic S. Natural history of multiple sclerosis: a unifying concept. Brain 2006;129:606–616.

8. Scalfari A, Neuhaus A, Degenhardt A, et al. The natural history of multiple sclerosis: a geographically based study 10: relapses and long-term disability. Brain : a journal of neurology 2010;133:1914–1929.

9. Klineova S, Lublin FD. Clinical Course of Multiple Sclerosis. Cold Spring Harb Perspect Med 2018;8:a028928.

10. Weinshenker BG, Bass B, Rice GPA, et al. The natural history of multiple sclerosis: a geographically based study: 2 predictive value of the early clinical course. Brain 1989;112:1419–1428.

11. Andravizou A, Dardiotis E, Artemiadis A, et al. Brain atrophy in multiple sclerosis: mechanisms, clinical relevance and treatment options. Autoimmunity Highlights 2019;10:7.

12. Chard D. Brain atrophy in clinically early relapsing-remitting multiple sclerosis. Brain 2002;125:327–337.

13. Cree BAC, Hollenbach JA, Bove R, et al. Silent progression in disease activity-free relapsing multiple sclerosis. Ann Neurol 2019;85:653–666.

14. Ziemssen T, Akgün K, Brück W. Molecular biomarkers in multiple sclerosis. J Neuroinflammation 2019;16:272.

15. Thompson AJ, Banwell BL, Barkhof F, et al. Diagnosis of multiple sclerosis: 2017 revisions of the McDonald criteria. Lancet Neurol 2018;17:162–173.

16. McDonald WI, Compston A, Edan G, et al. Recommended diagnostic criteria for multiple sclerosis: guidelines from the international panel on the diagnosis of multiple sclerosis. Ann Neurol 2001;50:121–127.

17. Tintoré M, Rovira A, Río J, et al. Baseline MRI predicts future attacks and disability in clinically isolated syndromes. Neurology 2006;67:968–972.

18. Teixeira M, Seabra M, Carvalho L, et al. Clinically isolated syndrome, oligoclonal bands and multiple sclerosis. Clin Exp Neuroimmunol 2020;11:33–39.

19. Jafari A, Babajani A, Rezaei-Tavirani M. Multiple sclerosis biomarker discoveries by proteomics and metabolomics approaches. Biomark Insights 2021;16:11772719211013352.

20. Assarsson E, Lundberg M, Holmquist G, et al. Homogenous 96-plex PEA immunoassay exhibiting high sensitivity, specificity, and excellent scalability. PLoS One 2014;9:e95192.

21. Hu W, Loh L, Patel H, et al. Analytical validation of a multivariate proteomic serum based assay for disease activity assessments in multiple sclerosis. Americas Committee for Treatment and Research in Multiple Sclerosis (ACTRIMS) 2021 Forum; 2021; Virtual.

22. Bove R, Chitnis T, Cree BA, et al. SUMMIT (Serially Unified Multicenter Multiple Sclerosis Investigation): creating a repository of deeply phenotyped contemporary multiple sclerosis cohorts. Mult Scler 2018;24:1485–1498.

23. Qureshi F, Hu W, Loh L, et al. Analytical validation of a multi-protein, serum-based assay for disease activity assessments in multiple sclerosis. medRxiv. Preprint posted online January 31, 2023. doi:10.1101/2022.05.23.22275201.

24. Robin X, Turck N, Hainard A, et al. pROC: an open-source package for R and S+ to analyze and compare ROC curves. BMC Bioinformatics 2011;12:77.

25. R Core Team. R: A language and environment for statistical computing. R Foundation for Statistical Computing [online]. Available at: https://www.R-project.org/. Accessed January 13, 2023.

26. Raschka S. MLxtend: providing machine learning and data science utilities and extensions to Python’s scientific computing stack. J Open Source Softw 2018;3:638.

27. Håkansson I, Tisell A, Cassel P, et al. Neurofilament light chain in cerebrospinal fluid and prediction of disease activity in clinically isolated syndrome and relapsing-remitting multiple sclerosis. Eur J Neurol 2017;24:703–712.

28. Disanto G, Barro C, Benkert P, et al. Serum Neurofilament light: A biomarker of neuronal damage in multiple sclerosis. Annals of neurology 2017;81:857–870.

29. Novakova L, Axelsson M, Malmeström C, et al. NFL and CXCL13 may reveal disease activity in clinically and radiologically stable MS. Mult Scler Relat Disord 2020;46:102463.

30. Rosso M, Gonzalez CT, Healy BC, et al. Temporal association of sNfL and gad-enhancing lesions in multiple sclerosis. Ann Clin Transl Neurol 2020;7:945–955.

31. Matute-Blanch C, Villar LM, Álvarez-Cermeño JC, et al. Neurofilament light chain and oligoclonal bands are prognostic biomarkers in radiologically isolated syndrome. Brain 2018;141:1085–1093.

32. Barro C, Benkert P, Disanto G, et al. Serum neurofilament as a predictor of disease worsening and brain and spinal cord atrophy in multiple sclerosis. Brain 2018;141:2382–2391.

33. Chitnis T, Gonzalez C, Healy BC, et al. Neurofilament light chain serum levels correlate with 10-year MRI outcomes in multiple sclerosis. Ann Clin Transl Neurol 2018;5:1478–1491.

34. Novakova L, Axelsson M, Khademi M, et al. Cerebrospinal fluid biomarkers as a measure of disease activity and treatment efficacy in relapsing-remitting multiple sclerosis. J Neurochem 2017;141:296–304.

35. Manouchehrinia A, Stridh P, Khademi M, et al. Plasma neurofilament light levels are associated with risk of disability in multiple sclerosis. Neurology 2020;94:e2457–e2467.

36. Kuhle J, Disanto G, Lorscheider J, et al. Fingolimod and CSF neurofilament light chain levels in relapsing-remitting multiple sclerosis. Neurology 2015;84:1639–1643.

37. Kuhle J, Plavina T, Barro C, et al. Neurofilament light levels are associated with long-term outcomes in multiple sclerosis. Mult Scler 2020;26:1691–1699.

38. Zetterberg H. Neurofilament Light: A Dynamic Cross-Disease Fluid Biomarker for Neurodegeneration. Neuron 2016;91:1–3.

39. Gaetani L, Blennow K, Calabresi P, Di Filippo M, Parnetti L, Zetterberg H. Neurofilament light chain as a biomarker in neurological disorders. Journal of Neurology, Neurosurgery &amp; Psychiatry 2019;90:870–881.

40. Barro C, Chitnis T, Weiner HL. Blood neurofilament light: a critical review of its application to neurologic disease. Ann Clin Transl Neurol 2020;7:2508–2523.

41. Graber JJ, Ford D, Zhan M, Francis G, Panitch H, Dhib-Jalbut S. Cytokine changes during interferon-beta therapy in multiple sclerosis: correlations with interferon dose and MRI response. J Neuroimmunol 2007;185:168–174.

42. Comabella M, Balashov K, Issazadeh S, Smith D, Weiner HL, Khoury SJ. Elevated interleukin-12 in progressive multiple sclerosis correlates with disease activity and is normalized by pulse cyclophosphamide therapy. J Clin Invest 1998;102:671–678.

43. Brettschneider J, Czerwoniak A, Senel M, et al. The chemokine CXCL13 is a prognostic marker in clinically isolated syndrome (CIS). PLoS One 2010;5:e11986–e11986.

44. DiSano KD, Gilli F, Pachner AR. Intrathecally produced CXCL13: A predictive biomarker in multiple sclerosis. Mult Scler J Exp Transl Clin 2020;6:2055217320981396-2055217320981396.

45. Khademi M, Kockum I, Andersson ML, et al. Cerebrospinal fluid CXCL13 in multiple sclerosis: a suggestive prognostic marker for the disease course. Mult Scler 2011;17:335–343.

46. Stilund M, Gjelstrup MC, Petersen T, Møller HJ, Rasmussen PV, Christensen T. Biomarkers of inflammation and axonal degeneration/damage in patients with newly diagnosed multiple sclerosis: contributions of the soluble CD163 CSF/serum ratio to a biomarker panel. PLoS One 2015;10:e0119681.

47. Miller JR. The importance of early diagnosis of multiple sclerosis. J Manag Care Pharm 2004;10:S4–S11.

48. Kaunzner UW, Gauthier SA. MRI in the assessment and monitoring of multiple sclerosis: an update on best practice. Ther Adv Neurol Disord 2017;10:247–261.

49. Chitnis T, Foley J, Ionete C, et al. Multivariate Proteomic MS Disease Activity Test Result Distributions Based on Disease Modifying Therapy Categories. 7th Annual Americas Committee for Treatment and Research in Multiple Sclerosis (ACTRIMS) Forum; 2022 February 24-26, 2022; West Palm Beach, Florida.

50. Paul A, Comabella M, Gandhi R. Biomarkers in Multiple Sclerosis. Cold Spring Harb Perspect Med 2019;9.

